# Cell-free RNA Signatures Derived from the Tumor Microenvironment Predict Outcomes of CAR-T Therapy in Large B Cell Lymphoma

**DOI:** 10.64898/2026.03.16.26348550

**Authors:** Conor Loy, Glory Agun, Katie Maurer, Anna Bosch Vilaseca, Daria Potapova, Caron A. Jacobson, Jerome Ritz, Iwijn De Vlaminck

## Abstract

Anti-CD19 chimeric antigen receptor (CAR) T-cell therapy can induce durable remissions in patients with large B-cell lymphoma (LBCL), yet outcomes remain variable. Reliable pre-treatment predictors of durable response remain limited, leaving a critical gap in patient management. To address this, we profiled pre-treatment plasma cell-free RNA (cfRNA) from 91 LBCL patients treated with axicabtagene ciloleucel (axi-cel, Yescarta) across three independent cohorts. We first demonstrated that signatures of “lymph node-like” tumor microenvironments (TMEs), previously identified in tumor biopsies and shown to correlate with favorable outcomes, are specifically elevated in the pre-treatment plasma cfRNA of responders, but not in matched peripheral blood mononuclear cells (PBMCs). These observations indicate that cfRNA captures TME tissue-derived signals not reflected in circulating immune cells. Next, using unbiased approaches, we identified additional cfRNA signatures associated with one-year clinical outcomes that capture the underlying biological landscape of treatment response. Collectively, these findings support pre-treatment plasma cfRNA as a minimally invasive surrogate of TME state to prospectively inform durable CAR T-cell therapy outcomes and guide risk stratification and TME-modulating adjunct therapies.

## INTRODUCTION

Anti-CD19 chimeric antigen receptor (CAR) T cell therapy has transformed care for patients with relapsed or refractory Large B-cell Lymphoma (LBCL). However, despite high initial response rates, nearly half of patients experience primary resistance or relapse within one year of treatment (1,2). While post-infusion factors, such as CAR T-cell expansion and persistence influence efficacy, the baseline, pre-treatment tumor state is increasingly recognized as a key determinant of treatment outcomes (3–6). Identifying pre-treatment determinants is essential for patient stratification and the development of targeted adjunct therapies.

The tumor microenvironment (TME) has emerged as a critical regulator of CAR T-cell outcomes in LBCL (3,7,8). Recent work by Li et al. defined three TME archetypes using transcriptomic profiling of pre-treatment tumor biopsies: lymph node–like (LN), follicular macrophage and accessory cell (FMAC), and T-cell exhausted (TEX). Patients with LN-like TMEs exhibit improved response rates, whereas FMAC- or TEX-dominant microenvironments are linked to higher rates of therapeutic failure. Beyond the TME, pre-treatment systemic immune features have also been associated with therapy outcomes. Maurer et al. demonstrated that circulating B cell abundance prior to leukapheresis correlates with CAR T-cell response (9). While circulating immune cell counts can be easily measured, direct characterization of the TME requires invasive tissue biopsies that are not routinely performed and carry procedure-related risk (3).

Cell-free RNA (cfRNA) in plasma is a minimally invasive biomarker shed by cells in tissues and the circulation (10). Unlike DNA-based liquid biopsies that primarily focus on mutational burden, cfRNA reflects the real-time transcriptional activity of both circulating and tissue-resident cells (11). Because cfRNA integrates signals from these multiple compartments, it may provide a systems-level, tissue-informed readout of disease biology(12–15). Given the unique biological origin of cfRNA, we tested the potential of plasma cfRNA as a minimally invasive, accessible biomarker for characterizing TME activity and predicting response to CAR T therapy. We assayed pre-treatment plasma from patients with LBCL receiving anti-CD19 CAR T cell therapy across three independent patient cohorts. To evaluate the tissue-specificity of these signals, we integrated matched single cell peripheral blood mononuclear cell (PBMC) transcriptomes and compared our findings against published gene signatures derived from tumor biopsies.

We demonstrate that the LN-like archetype signature is recapitulated in plasma cfRNA, but is absent from PBMC transcriptomes, indicating that cfRNA captures immune processes within the TME that are not captured by circulating cells. Machine learning models leveraging these cfRNA signatures achieved a median test AUC of 0.73, significantly outperforming models based on other feature sets. Furthermore, using unbiased profiling of the cfRNA transcriptomes, we identified gene signatures, including transcripts such as *LTB*, *SPDYC*, and *IGHD*, that were predictive of one-year clinical response. Collectively, these findings support pre-infusion cfRNA profiling as an informative and minimally invasive liquid-biopsy approach for durable response prediction, and biological stratification of patients prior to therapy.

## RESULTS

### Clinical cohort and cfRNA profiling

We analyzed pre-treatment plasma samples from 91 patients with relapsed or refractory LBCL receiving axicabtagene ciloleucel (axi-cel) at the Dana-Farber Cancer Institute across three independent cohorts (Cohort 1, n=28; Cohort 2, n=28; Cohort 3, n=35; **Table S1, Figure 1A**). The median age at infusion was 65 years (range 24-83), and the majority of patients had *de novo* DLBCL (74%) and stage III-IV disease (86%). Samples were collected prior to treatment, either near the time of leukapheresis or before lymphodepletion. Responders were defined as patients with partial or complete response by PET imaging at one-year post-infusion, or at the latest follow-up for patients with less than one year of follow-up (**Figure 1B**). cfRNA sequencing was performed on EDTA-collected plasma to a median depth of 41 million reads, identifying a median of 5 million unique features per sample (**Figure 1C-D, Methods**). Principal component analysis showed no clustering by collection timepoint or cohort, indicating minimal batch-specific biases (**Figure 1D, Methods**)

**Figure 1.**
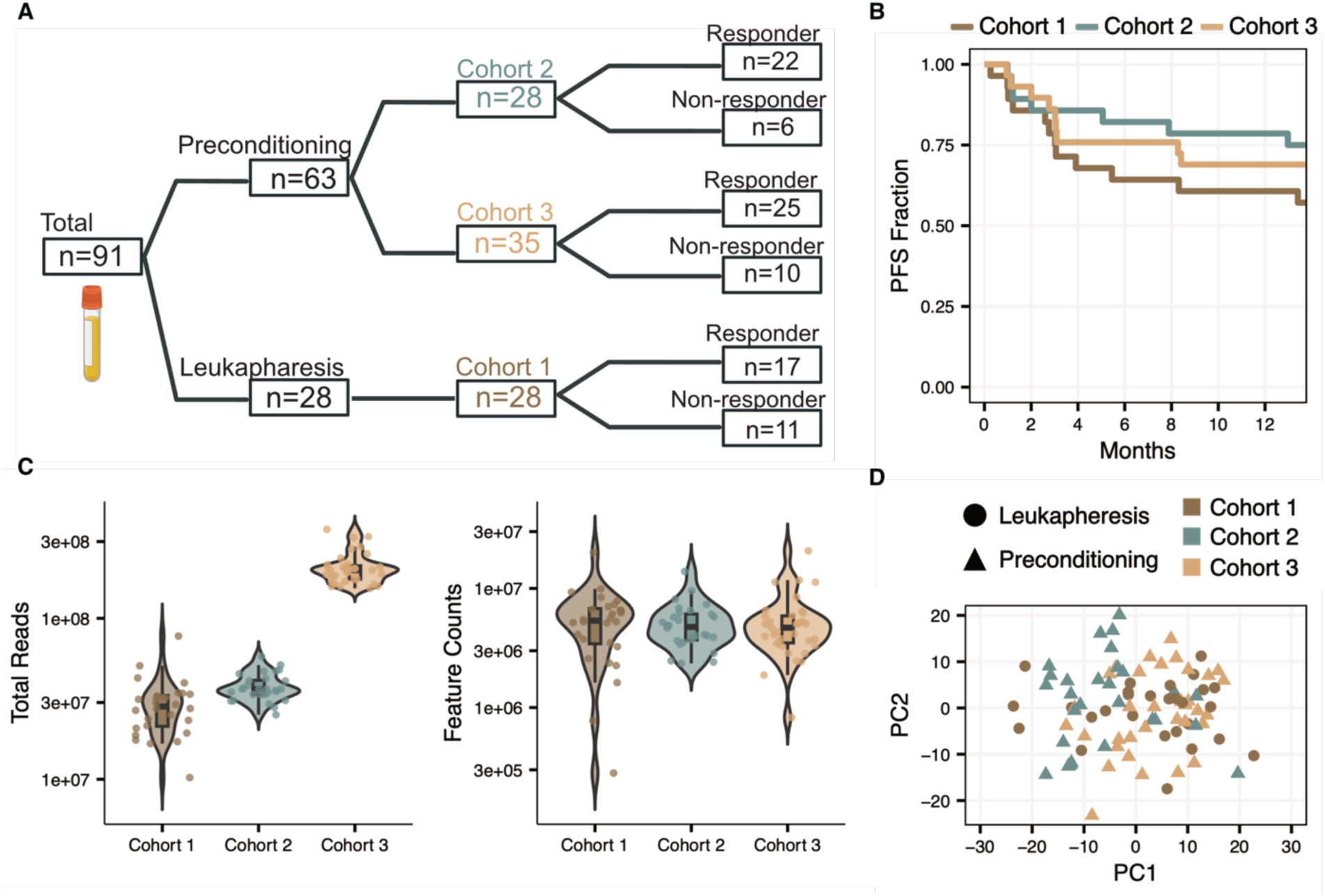
Cohort Overview. **A)** Sample cohort overview. **B)** Progression-free survival (PFS) curves stratified by cohort. **C)** Distributions of sequencing depth and unique counted features in each cohort. **D)** Principal component analysis (PCA) on the top 500 most variable transcripts. Cohort indicated by color and timepoint by shape.

### Signatures previously discovered in TME biopsies are recapitulated in cfRNA

We first investigated whether the TME archetype gene markers identified in tumor biopsies by Li et al. are differentially abundant in the cfRNA or PBMCs of responders and non-responders. To this end, we calculated marker gene module scores for each archetype (LN, FMAC, and TEX) across three compartments: TME biopsies, plasma cfRNA, and PBMCs (3,9). The TME biopsy data were sourced from Li et al., while the PBMC samples included a subset collected from the same blood draws as the cfRNA samples, providing a direct comparison.

Consistent with the findings by Li et al., responders exhibited higher LN module scores than non-responders within the TME biopsies (**Figure 2A**). Importantly, a similar signal was observed in the cfRNA data in each of the three patient cohorts, but this signal was absent in the matched PBMCs. This observation indicates that the LN TME signature detected in plasma reflects tissue-derived transcriptional activity rather than differences in circulating immune cell composition. Furthermore, patients with above-median cfRNA LN scores demonstrated significantly improved progression-free survival (PFS) across all three cohorts (**Figure 2B**; Wald test, p = 0.01). To confirm that this signal did not originate from specific circulating populations, we compared LN-module scores across individual PBMC cell types; we found no significant differences between responders and non-responders in any cellular subset (**Figure S1**). In contrast to the LN archetype, no significant differences were observed in cfRNA or PBMCs for FMAC or TEX module scores (**Figure S2A**). Additionally, we applied the LymphoMap algorithm defined by Li et al. on the cfRNA-seq data and found no difference in PFS between the labelled sample groups, likely due to the algorithm’s reliance on transcript weights optimized specifically for solid tissue biopsies (**Figure S2B**).

**Figure 2.**
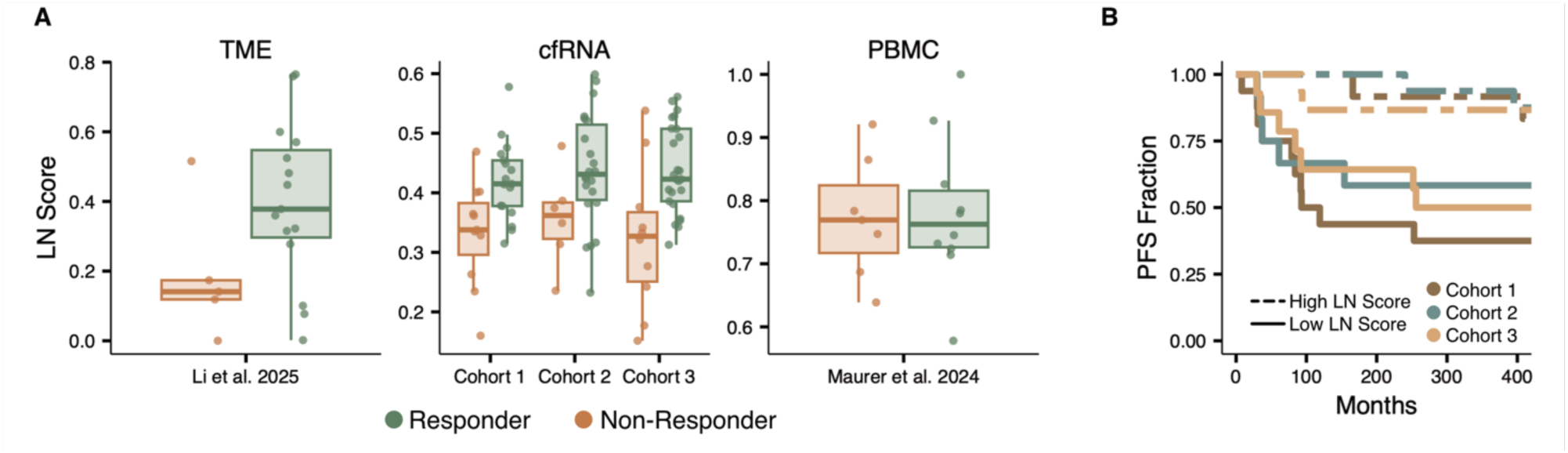
LN Module Score. **A)** LN archetype module scores in responders and non-responders across three data sources: TME biopsies (Li et al. 2025), plasma cfRNA (cohorts 1-3), and PBMCs (Maurer et al. 2024). Module scores were calculated using expression-level-controlled background subtraction (**Methods**). **B)** Progression-free survival stratified by above median (high) and below-median (low) cfRNA LN module scores across all three cohorts.

Collectively, these results demonstrate that TME archetypes can be characterized, and patient outcomes predicted, from a standard blood draw prior to treatment using cfRNA profiling. This highlights the utility of plasma cfRNA as a minimally invasive window into disease biology previously accessible only through solid tissue biopsies.

### cfRNA predictive performance

We next evaluated the ability of cfRNA to predict treatment response using Monte Carlo cross-validation across four distinct feature sets: the full cfRNA transcriptome, and the individual LN, FMAC, and TEX archetype marker sets. For each feature set, we performed 101 training iterations and recorded the selected model features, feature weights, and test performance as measured by the area under the receiver operating characteristic curve (AUC) (**Methods**).

Models leveraging LN archetype marker transcripts achieved the highest predictive performance, with a median test AUC of 0.73, significantly outperforming all other feature sets (**Figure 3A**). Models using the full transcriptome or FMAC archetype marker transcripts showed intermediate performance (median AUC, all transcripts = 61.25 and FMAC = 65), while those using TEX archetype marker transcripts performed poorly (median AUC=52.5). These findings indicate that while biopsy-informed feature selection (LN) yields the highest performance, the FMAC archetype also contains non-random predictive capacity, and a biopsy-naïve approach across the entire transcriptome can independently recover predictive signals.

**Figure 3.**
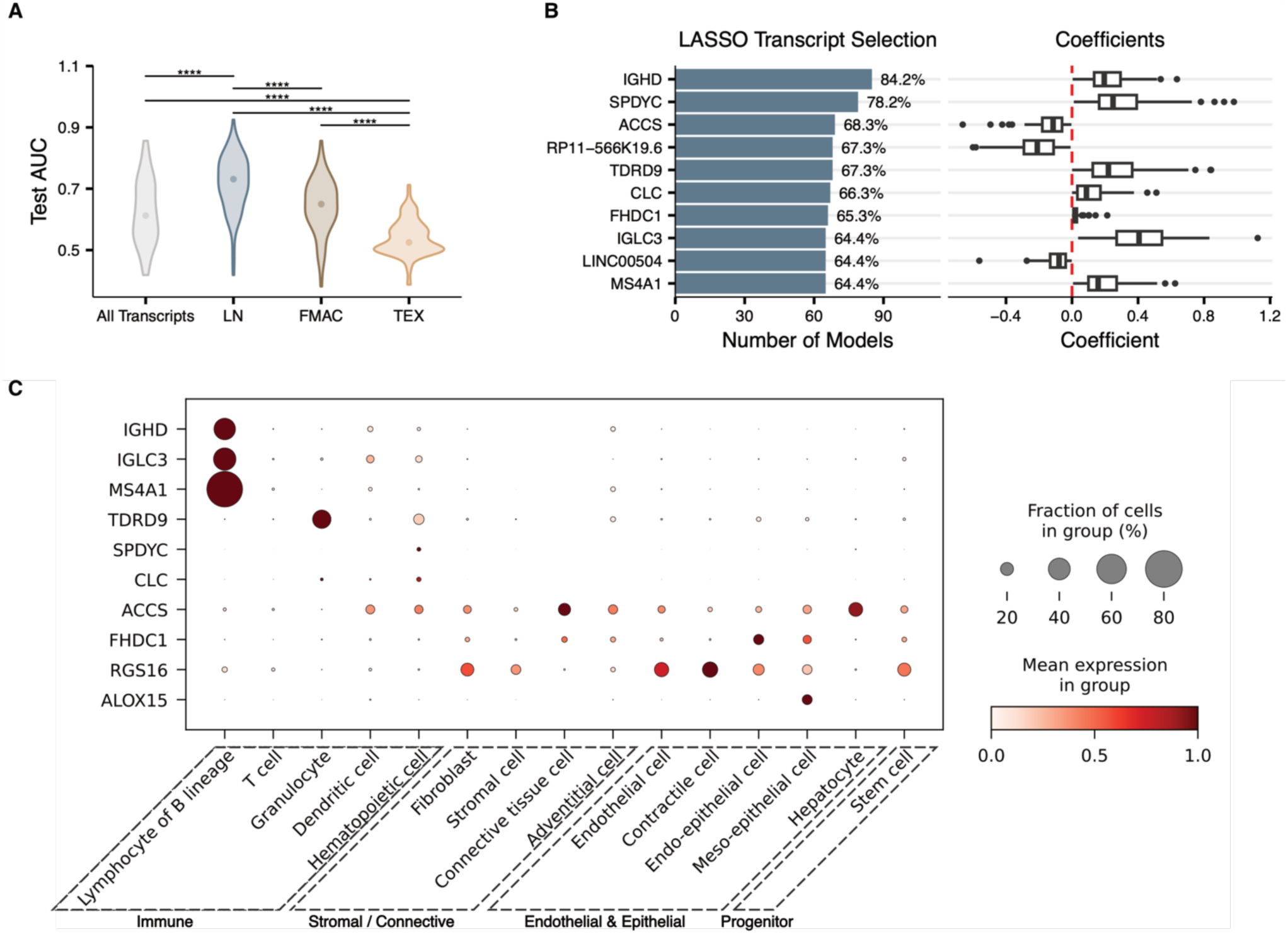
cfRNA predictive signatures and their origins. **A)** Distributions of test-set AUC values across 101 Monte Carlo cross-validation iterations for LASSO models trained on four cfRNA feature sets: all transcripts, LN archetype markers, FMAC archetype markers, and TEX archetype markers. Significance assessed by Mann-Whitney U test (****p < 0.0001). **B)** Top 10 most frequently selected transcripts across 101 LASSO iterations using all transcripts as input features. Bar plot (left) shows selection frequency; box plot (right) shows corresponding model coefficients across iterations. Red dashed line indicates zero. **C)** Dot plot showing mean expression (color) and fraction of expressing cells (size) for the top selected protein-coding transcripts across cell types from the Tabula Sapiens cell atlas, grouped by broad lineage category.

Analysis of the transcripts most frequently selected in the all-transcript models identified *IGHD* (84.2% of models) and *SPDYC* (78.2%) as the top features (**Figure 3B**). To resolve the cellular origins of these transcripts, we mapped the top protein-coding transcripts to the Tabula Sapiens cell atlas (**Figure 3C**) (16). While *IGHD*, *IGLC3*, and *MS4A1* (CD20) were predominantly expressed in B cell lineages, *ACCS*, *RGS16*, and *FHDC1* showed broad expression across stromal, endothelial, and epithelial compartments, indicating that cfRNA captures predictive signals from multiple cellular sources.

Several of these top features have established roles in lymphoma biology and immunotherapy response. *IGHD*, *IGLC3*, and *MS4A1* are B cell lineage markers whose abundance likely reflects tumor burden or the presence of tumor-resident B cells (17). *RGS16*, a regulator of G-protein signaling involved in chemokine receptor signaling, is implicated in T cell trafficking and immune cell recruitment to the TME (18,19). *ACCS* showed expression predominantly in stromal and endothelial compartments, suggesting a non-immune TME contribution to the model. *SPDYC*, a speedy cell cycle regulator homolog, is poorly characterized in lymphoma and warrants further investigation.

Collectively, these results demonstrate that cfRNA captures multiple axes of TME biology, spanning immune, stromal, and endothelial compartments, that are predictive of treatment response from a single blood draw. Together with the LN-restricted modeling results, these data suggest that integrating biologically informed feature sets with unbiased transcriptome-wide discovery may yield the most robust predictive framework as larger datasets become available.

### Signatures discovered in cfRNA are recapitulated in TME biopsies

Having identified predictive features through joint selection with LASSO regression, we next performed differential abundance analysis to characterize the broader transcriptomic differences between responders and non-responders through independent transcript modeling (20). To ensure robust discovery, we used cohorts 1 and 2 for discovery and reserved Cohort 3 as an independent validation set (**Methods**).

We identified 776 differentially abundant transcripts between responders and non-responders (DESeq2, Benjamini-Hochberg (BH) adjusted p-value < 0.1, **Figure 4A**). The log_2_ fold changes of these transcripts were highly correlated with those observed in cohort 3 (Pearson r=0.8, **Figure S3**), confirming the reproducibility of the cfRNA signature across distinct patient groups. Among the top differentially abundant transcripts were Lymphotoxin Beta (*LTB*) and Killer Cell Lectin-Like Receptor B1 (*KLRB1*), both LN archetype marker genes, as well as Cadherin 5 (*CDH5*) and Divergent Protein Kinase 2B (*DIPK2B*), both FMAC archetype marker genes (**Figure S4**). Notably, no TEX archetype marker genes used in computing module scores were detected, consistent with the poor predictive performance of TEX features in the LASSO models (**Figure 3A**).

**Figure 4.**
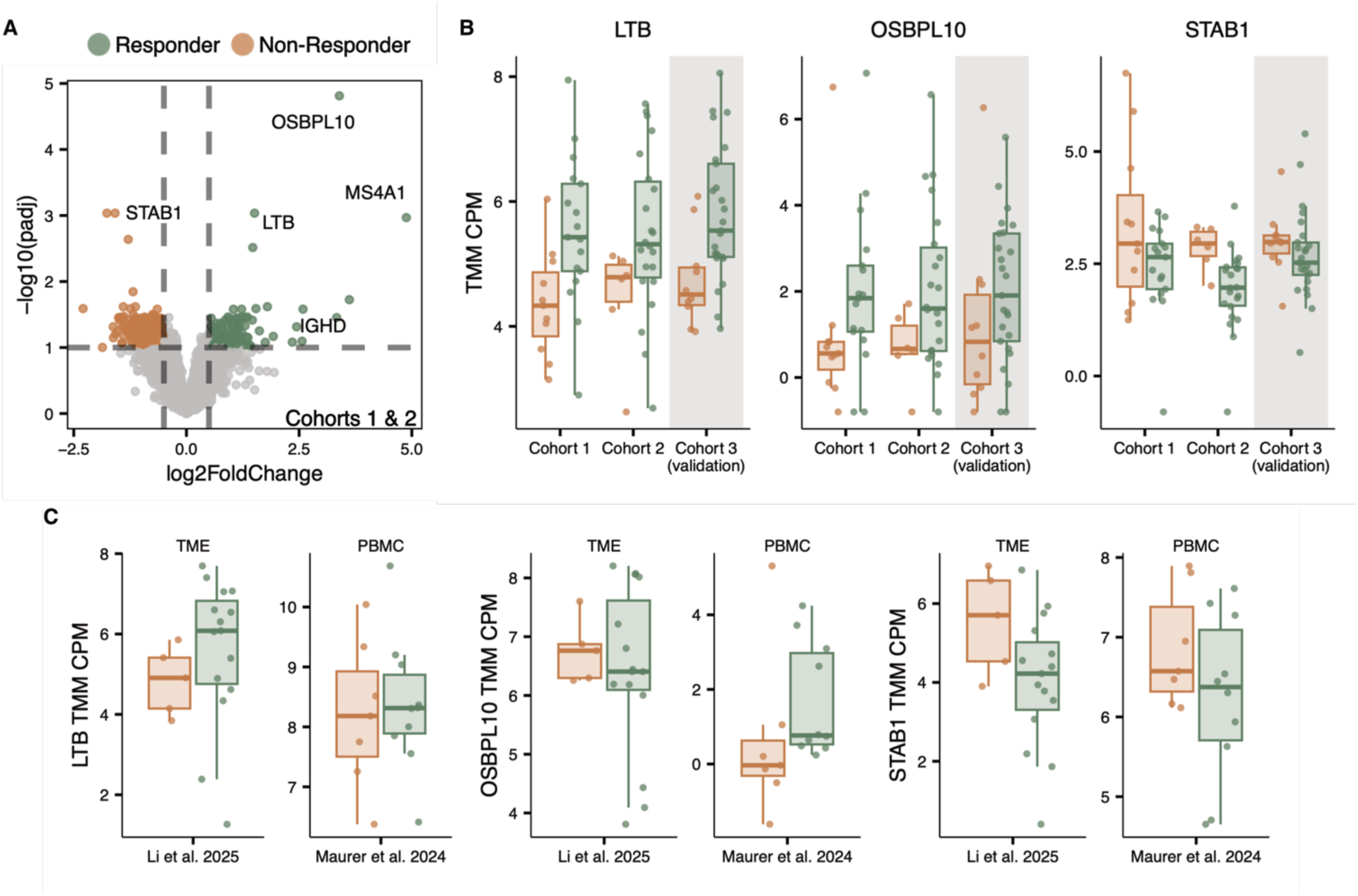
Differential abundance analysis identifies cfRNA biomarkers of durable response. **A)** Volcano plot of differentially abundant transcripts between responders (green) and non-responders (orange) in the discovery cohorts (Cohorts 1 and 2; DESeq2, BH-adjusted p < 0.1). **B)** Normalized cfRNA abundance (log2 TMM CPM) of *LTB*, *OSBPL10*, and *STAB1* across the discovery cohorts (Cohorts 1 and 2) and the validation cohort (Cohort 3) **C)** Pseudobulk expression of *LTB*, *OSBPL10*, and *STAB1* in TME biopsies (Li et al. 2025) and PBMCs (Maurer et al. 2024), comparing responders and non-responders.

In addition to TME archetype-associated transcripts, we identified novel transcripts associated with durable response (**Figure 4B**). Oxysterol-binding protein-like 10 (*OSBPL10*), a lipid transporter implicated in tumorigenesis, was elevated in patients with durable response (21). Conversely, stabilin-1 (*STAB1*) abundance was lower in responders. *STAB1* encodes a transmembrane receptor expressed in M2-like macrophages that promote an immunosuppressive TME (22).

To contextualize these findings, we examined pseudobulk expression patterns of selected transcripts in TME biopsies and PBMC datasets. Responders exhibited elevated *LTB* expression within the TME, but not in PBMCs (**Figure 4C**), supporting a tissue-specific origin. In contrast, *OSBPL10* expression patterns were recapitulated in PBMCs but not in TMEs, whereas *STAB1* showed similar directional changes across both TME and PBMC compartments (**Figure 4C**). These compartment-specific patterns suggest that cfRNA integrates both local microenvironmental signals and systemic immune features, with distinct transcripts reflecting different biological sources.

These findings demonstrate that unbiased differential abundance analysis independently recovers TME-associated biology while also revealing systemic immune signatures. The concordance between cfRNA-derived signatures and tumor biopsy data provides reciprocal validation that plasma cfRNA reflects biologically meaningful microenvironmental programs relevant to CAR T-cell response.

## DISCUSSION

We demonstrate that pre-infusion plasma cfRNA signatures predict response to anti-CD19 CAR T-cell therapy by capturing the immunological state of the tumor microenvironment (TME). A key observation of our study is the bidirectional recapitulation of these signals: signatures previously discovered in TME biopsies are present in plasma cfRNA, and conversely, signatures independently developed from the plasma cell-free transcriptome are mirrored in the underlying tumor tissue. Specifically, we found that cfRNA signatures reflecting lymphoid structure and fibrosis serve as predictors of CAR T response. Within this framework, a lymph node-like (LN) TME phenotype, consistent with an immunologically active microenvironment, emerges as a positive predictor of durable therapeutic success. A second key observation is that these predictive TME features are notably absent in matched peripheral blood mononuclear cells (PBMCs), supporting a tissue-derived origin rather than a reflection of circulating immune composition.

Our analysis independently identified individual transcripts with direct relevance to TME biology and mechanisms of treatment resistance. We discovered that *LTB* cfRNA transcripts were elevated in responders. *LTB*, a type II membrane protein in the TNF superfamily associated with lymphoid tissue organization and immune activation, was also identified by Li et al as a marker gene for the LN archetype (3,23,24). Conversely, we also discovered *STAB1* cfRNA transcripts were elevated in non-responders. *STAB1* encodes stabilin-1, a scavenger receptor expressed on immunosuppressive tumor-associated macrophages associated with T cell exclusion and poor prognosis (25). Together, these findings indicate that cfRNA captures both structural features of organized antitumor immunity and signatures of myeloid-mediated immune suppression. By integrating PBMC and TME single-cell data, we found that while *LTB* transcripts likely originate from the TME and markers like *OSBPL10* appear to be systemic, indicating that cfRNA provides a dual window into local microenvironment state and systemic immune features.

The TME is known to regulate treatment response beyond CAR T-cell therapy, including in the setting of checkpoint inhibitors, bispecific antibodies, and antibody-drug conjugates (26). However, routine TME characterization has been limited by the need for invasive tissue biopsies. A blood-based readout of TME state could inform treatment selection and monitoring across immuno-oncology settings where tissue access is limited or serial sampling is needed. This is particularly relevant as TME-modulating therapies emerge as rational adjuncts to immunotherapies, including focal radiotherapy, small-molecule immunomodulation, myeloid-targeting agents, and microbiome-directed interventions (27–31). A plasma-based measure of TME activity could serve a dual role: identifying patients with unfavorable TME profiles who may benefit from pre-infusion conditioning and providing a rapid readout to verify TME remodeling before proceeding to CAR T-cell infusion.

This study has several limitations. The cfRNA sample set analyzed, while spanning three independent patient groups and collections, is moderate in size and drawn from a single institution. Additionally, cfRNA composition may be influenced by tumor burden, disease stage, and other clinical variables that were not fully controlled for in the current analysis. Future studies should evaluate cfRNA-based TME profiling alongside established clinical covariates and genomic biomarkers in larger, multi-institutional cohorts.

These findings motivate prospective evaluation of cfRNA-guided risk stratification before CAR T-cell infusion, extension to additional cell therapy products and disease settings, and integration with TME-modulating treatment strategies. If validated, plasma cfRNA profiling may enable scalable, minimally invasive assessment of microenvironmental state to support patient stratification and therapeutic development in LBCL and potentially other malignancies.

## MATERIALS AND METHODS

### Ethics Statement

The study was approved by the Dana-Farber/Harvard Cancer Center’s Office of Human Research. All recipients provided written informed consent.

### Clinical Cohort

Peripheral blood was collected from patients with LBCL undergoing axicabtagene ciloleucel (axi-cel; YESCARTA®) therapy at DFCI. Samples were obtained at leukaphereis or prior to pre-conditioning therapy. Disease status was evaluated by ^18F-FDG PET/CT at 12 months after infusion and patients were classified as responders (complete response or partial response) or non-responders (stable or progressive disease).

### Sample Collection

Blood samples were collected through standard venipuncture in EDTA tubes. Plasma was extracted through blood centrifugation and stored in 1-to 2-mL aliquots at −80 °C. Plasma samples were shipped from the Dana-Farber Cancer Institute to Cornell University on dry ice.

### Sample Processing and Sequencing

Plasma aliquots (1-2mL) were thawed at room temperature and centrifuged at 1,300 × g for 10 min at 4 °C. cfRNA was purified from the supernatant with the Plasma/Serum Circulating and Exosomal RNA Purification Mini Kit (cat. 51000, Norgen Biotek) following the manufacturer’s instructions. Eluates (14 µL) were treated with 3 µL TURBO DNase mix (AM2238, Invitrogen) and 1 µL Baseline-ZERO DNase (DB0715K, Lucigen) for 30 min at 37 °C, then cleaned and concentrated to 12 µL with the RNA Clean & Concentrator-5 kit (R1015, Zymo Research). Libraries were prepared from 8 µL of cfRNA using the SMART-Seq Total RNA Pico Input with UMIs (ZapR Mammalian) preparation kit (634356, Takara Bio). The workflow employed random-primed reverse transcription, UMI tagging, rRNA depletion (ZapR), PCR amplification, and dual indexing with the SMART-Seq Unique Dual Index Kit (634752, Takara Bio). Library concentration was measured on a Qubit 3.0 Fluorometer with the dsDNA HS Assay Kit (Q32854, Invitrogen); fragment size distribution was assessed on a Fragment Analyzer 5200 with the HS NGS Fragment kit (DNF-474-0500, Agilent). Equimolar-pooled libraries were sequenced on an Illumina NovaSeq X platform to generate 150-bp paired-end reads.

### Bioinformatic processing

Raw FASTQ files were analyzed with an in-house Snakemake workflow (v 7.7.0)(32). Adapters and low-quality bases were removed with BBDuk(33). Trimmed reads were aligned with STAR (v 2.7.0f) to a composite reference consisting of the GENCODE GRCh38 primary assembly. Alignment files were deduplicated on UMI and mapping coordinates with UMI-tools(34). Gene-level counts were generated with featureCounts(35). Library sizes were normalized using TMM scaling factors and gene abundance was quantified as log2 counts per million.

### Sample quality filtering

Libraries were filtered on three criteria: DNA contamination, RNA degradation, and total feature counts. DNA contamination was calculated as the ratio of reads mapping to intronic versus exonic regions; samples with an intron-to-exon ratio > 3 were discarded. RNA degradation was assessed with the 5′/3′ coverage bias metric from Qualimap (v2.2.1)(36); samples with a bias more than three standard deviations above the cohort mean were excluded. Lastly, any library with < 100,000 total feature-aligned reads was removed.

### Single cell analysis

Publicly available single-cell RNA sequencing datasets from peripheral blood mononuclear cell (PBMC) samples and tumor microenvironment (TME) biopsies were analyzed. The PBMC samples were partially matched to the cfRNA samples analyzed and the data obtained from the Gene Expression Omnibus (accession number GSE273170). The TME biopsy dataset was accessed via CellxGene (https://cellxgene.cziscience.com/collections/4429f25e-5c91-4981-9cf2-6a1044c36732). The PBMC data were reprocessed and reclustered using the Leiden algorithm at a resolution of 0.4 to refine cell-type identification. For downstream comparisons, pseudobulk expression profiles were generated by summing counts within each sample and cluster. For cell-type specific analyses, clusters with less than 10 cells in more than 3 samples were excluded from the analyses. For module score analyses, pseudo-bulk library sizes were normalized using TMM scaling factors and gene abundance was quantified as log2 counts per million.

### Transcript module score calculation

To calculate archetype module scores we used the top 20 marker genes for each archetype (LN: *TNFRSF25, SELL, PIK3CD, IL16, BIRC3, CCL21, FOXP3, LY9, KLRB1, CD1C, LTB, CD40LG, LAMC2, SIGLEC5, MYBL1, ICOSLG, ATM, PDK1, C5, FCGR2B*; FMAC: *CLEC14 CD36, STC1, ANGPTL4, JAG1, PALMD, TIE1, ROBO4, SNAI1, ITGB3, CDH5, BCL6B, MMRN2, SPRY4, KDR, DIPK2B, JAG2, APLNR, TPM1, PDGFB*; TEX*: IDO1, GZMB, TLR8, GZMH, CCL4, CD244, LAG3, GZMA, CXCR6, FASLG, NKG7, CCR5, PRF1, SLAMF7, GZMK, CD8A, C1QA, C1QB, GBP1, FCGR1A*). Module scores were computed using an expression-level-controlled scoring approach adapted from Tirosh et al(37). For each gene set, the mean expression across member genes was calculated per sample. A background control score was derived from a set of genes randomly selected to match the expression-level distribution of the target genes (100 control genes per target gene, drawn from 25 expression bins). The module score was defined as the difference between the target and control means, then rescaled to the unit interval via min-max normalization.

### LymphoMap Algorithm

To classify samples based on predefined archetypes, the LymphoMapR algorithm was applied. The algorithm uses gene expression profiles to assign bulk RNA-seq samples to one of the three archetypes, LN, FMAC, TEX or Unassigned. Raw count data was used as input with the data type specified within function parameters. Classification was done using 50 features per archetype, a minimum log_2_ fold change of 1 and adjusted p-value of 0.05 to select features. Uniform Manifold Approximation and Projection (UMAP) embeddings were generated using inbuilt plotting function in package.

### Machine Learning

To evaluate the predictive performance of cfRNA features, we implemented Monte Carlo cross validation on multiple gene sets with 101 iterations. The top twenty marker transcripts for each archetype were used. In each iteration, samples were split into training (70%) and test (30%) sets, stratified by response status. A logistic regression model with L1 (LASSO) regularization was trained using five-fold cross-validation for hyperparameter optimization (scikit-learn, *liblinear* solver). Model performance was assessed by the area under the receiver operating characteristic curve (AUC) on the held-out test set. For each iteration, AUC values, predicted classifier scores, and non-zero-coefficient features were recorded to evaluate predictive stability and feature recurrence across splits.

### Single Cell Visualization

To examine the cellular origins of cfRNA transcripts most frequently selected by modeling, we analyzed single-cell RNA-sequencing data from the Tabula Sapiens v2 human cell atlas queried from CellxGene(16). Analysis was restricted to samples from healthy donors generated with the 10x 3′ v3 assay. Genes were limited to the most frequently selected mRNAs in the cfRNA modeling. Cell types were manually curated based on visual inspection of the data and relevance to the project. Scaled expression values were visualized using dot plots generated with Scanpy (*sc.pl.dotplot*), summarizing mean expression and detection frequency across the selected broad cell classes(38).

### Differential Abundance Analysis

Differential abundance analysis was performed using DESeq2(20) using samples from cohort 1 and cohort 2. Log2 fold changes between responders and non-responders was calculated separately using samples from cohort 3. Pearson correlation was used to compared fold change estimates from using cohort 1 and cohort 2 versus cohort 3.

### Statistical Analysis

Analyses were conducted in R and Python. Data processing and visualization used the R Tidyverse ecosystem(39), and machine learning was implemented with the Python scikit-learn, NumPy, and pandas libraries(40–42). Bioinformatic workflows were managed with Snakemake(32). Statistical comparisons were performed using Mann-Whitney U tests unless otherwise noted. Sequencing reads were aligned to the GRCh38 GENCODE v38 primary assembly, and gene-level counts were generated using the corresponding GENCODE v38 annotation(43).

## Data Availability

De-identified RNA-seq count matrices have been uploaded to the NCBI (National Center for Biotechnology Information) GEO (Gene Expression Omnibus) database and will be publicly available upon publication. All code has been deposited on GitHub and will be available upon publication.

## Acknowledgements

We thank the staff of the Pasquarello Tissue Bank in Hematologic Malignancies at the Dana-Farber Cancer Institute for providing plasma samples and the patients and their families for their help to further our understanding of large B-cell lymphoma.

## Supplementary Materials

**Figure S1.**
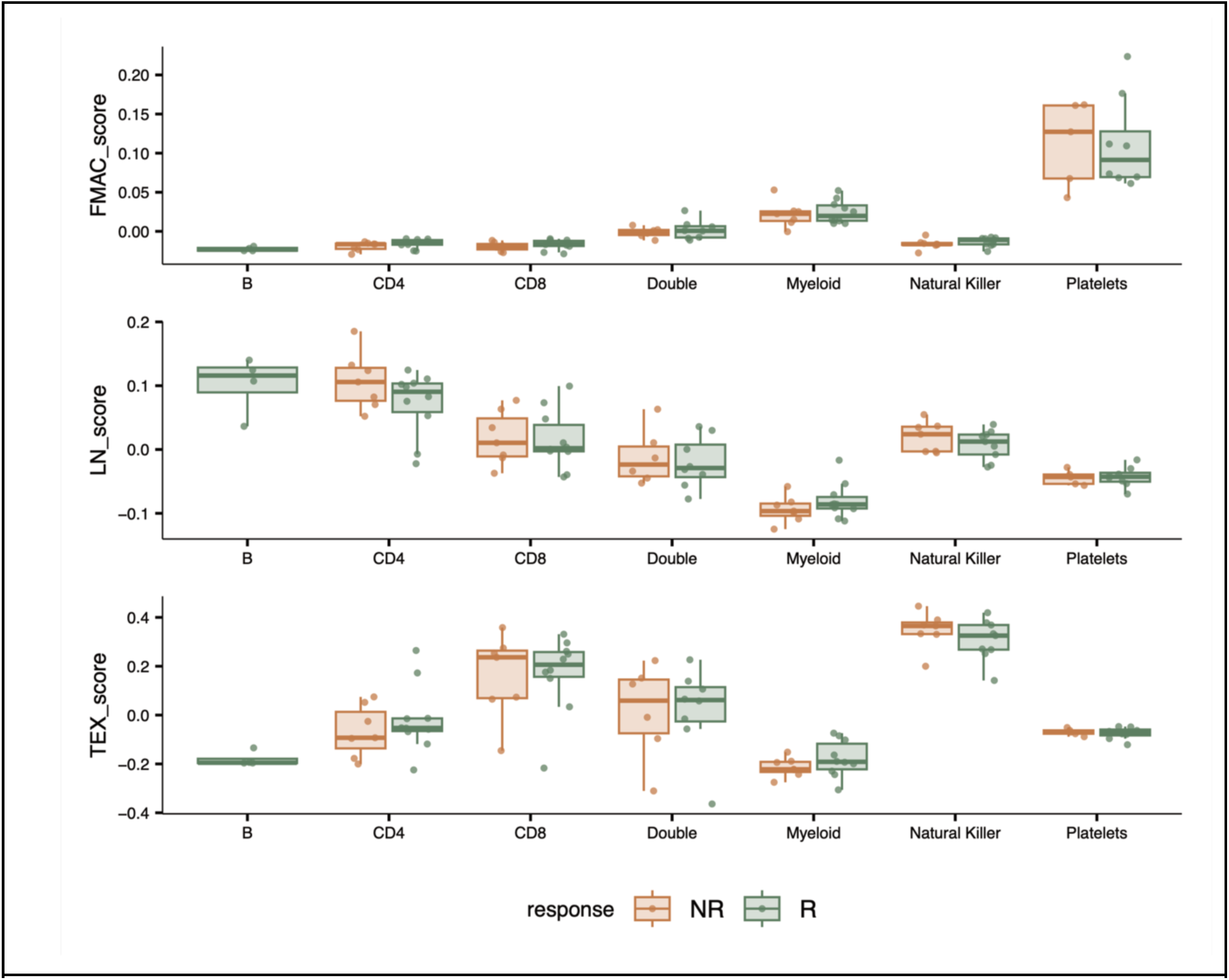
TME archetype module scores across PBMC cell types. FMAC (top), LN (middle), and TEX (bottom) archetype module scores computed from pseudobulk scRNA-seq profiles of major PBMC cell types, comparing responders (R, green) and non-responders (NR, orange). Sample-cell type pairings with less than 10 cells were removed from the analysis.

**Figure S2.**
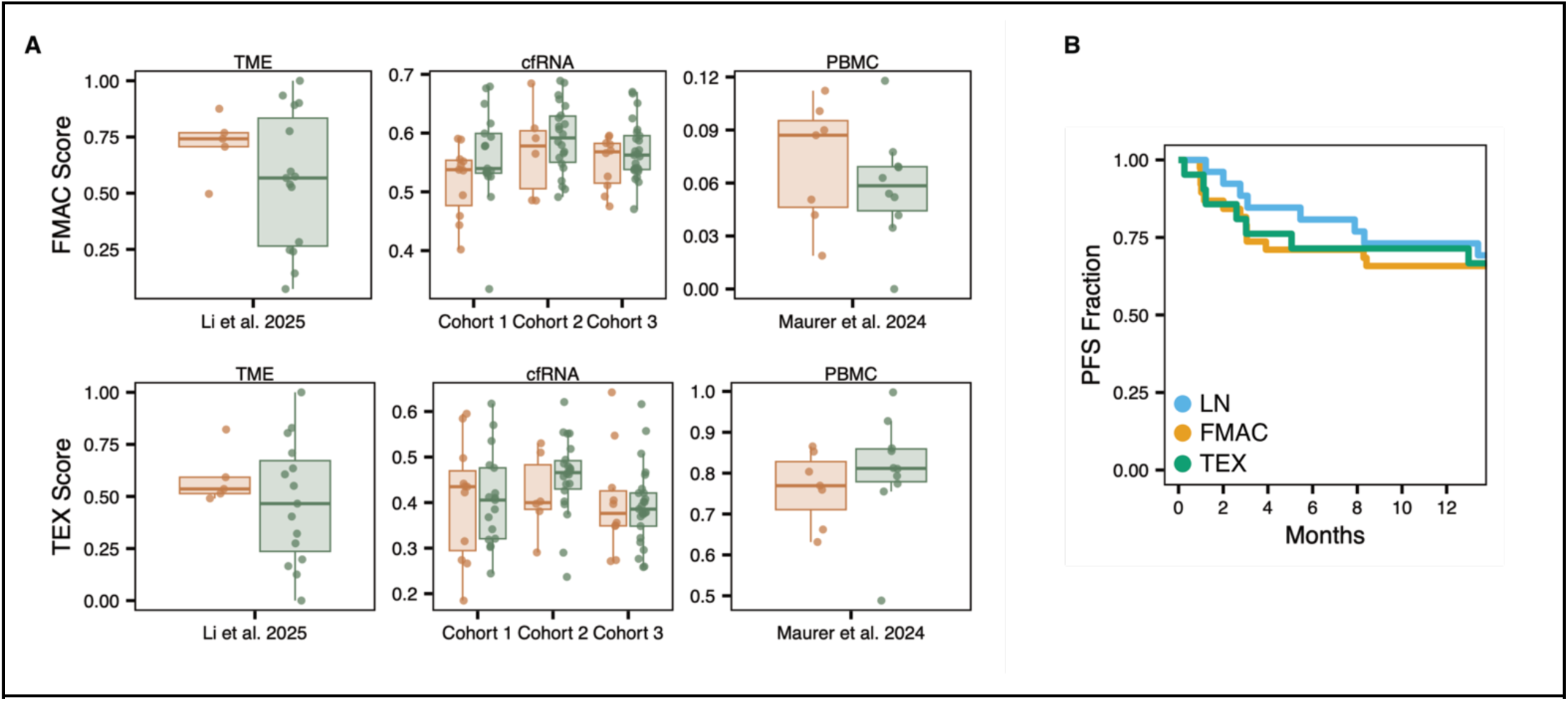
FMAC and TEX archetype scores and LymphoMap classification. **A)** FMAC (top) and TEX (bottom) archetype module scores in responders (green) and non-responders (orange) across TME biopsies (Li et al. 2025), plasma cfRNA (Cohorts 1-3), and PBMCs (Maurer et al. 2024). **B)** Progression-free survival curves for patients classified by the LymphoMap algorithm (Li et al. 2025) applied to cfRNA data. Patients were assigned to LN (blue), FMAC (orange), or TEX (green) groups.

**Figure S3.**
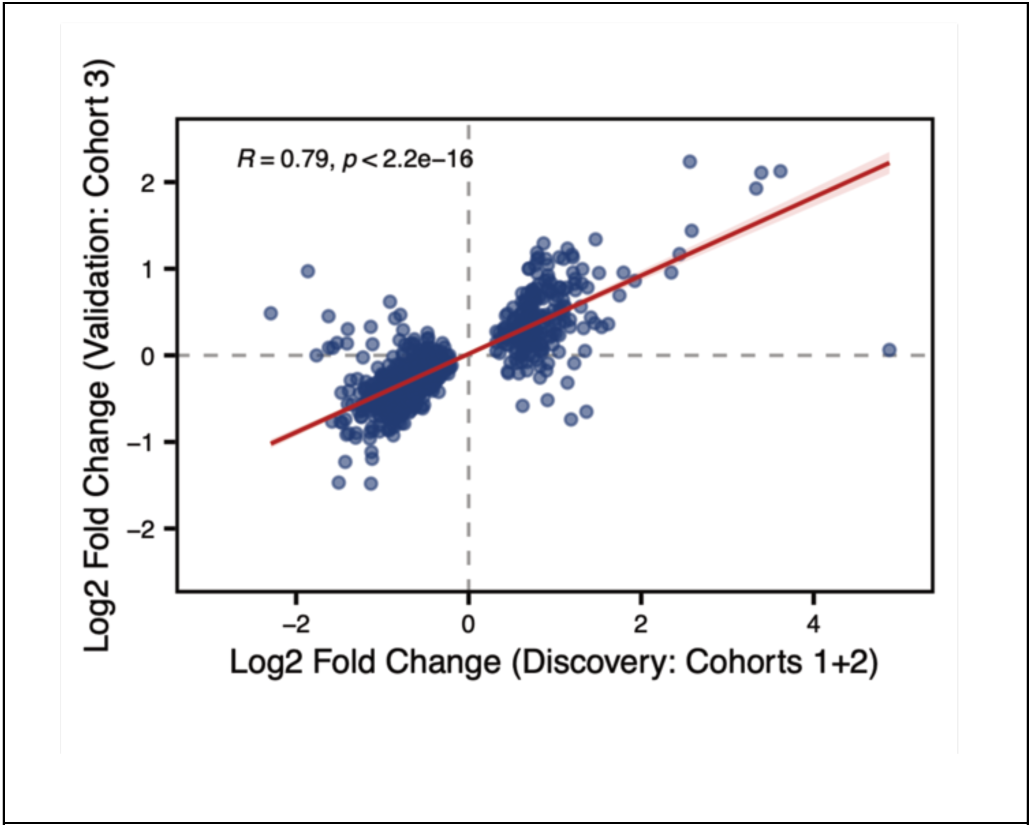
Correlation of log₂ fold changes for transcripts differentially abundant between responders and non-responders in the discovery cohort (Cohorts 1 and 2) compared with the independent validation cohort (Cohort 3). Red line indicates linear regression fit with 95% confidence interval (Pearson R = 0.79, p < 2.2e-16).

**Figure S4.**
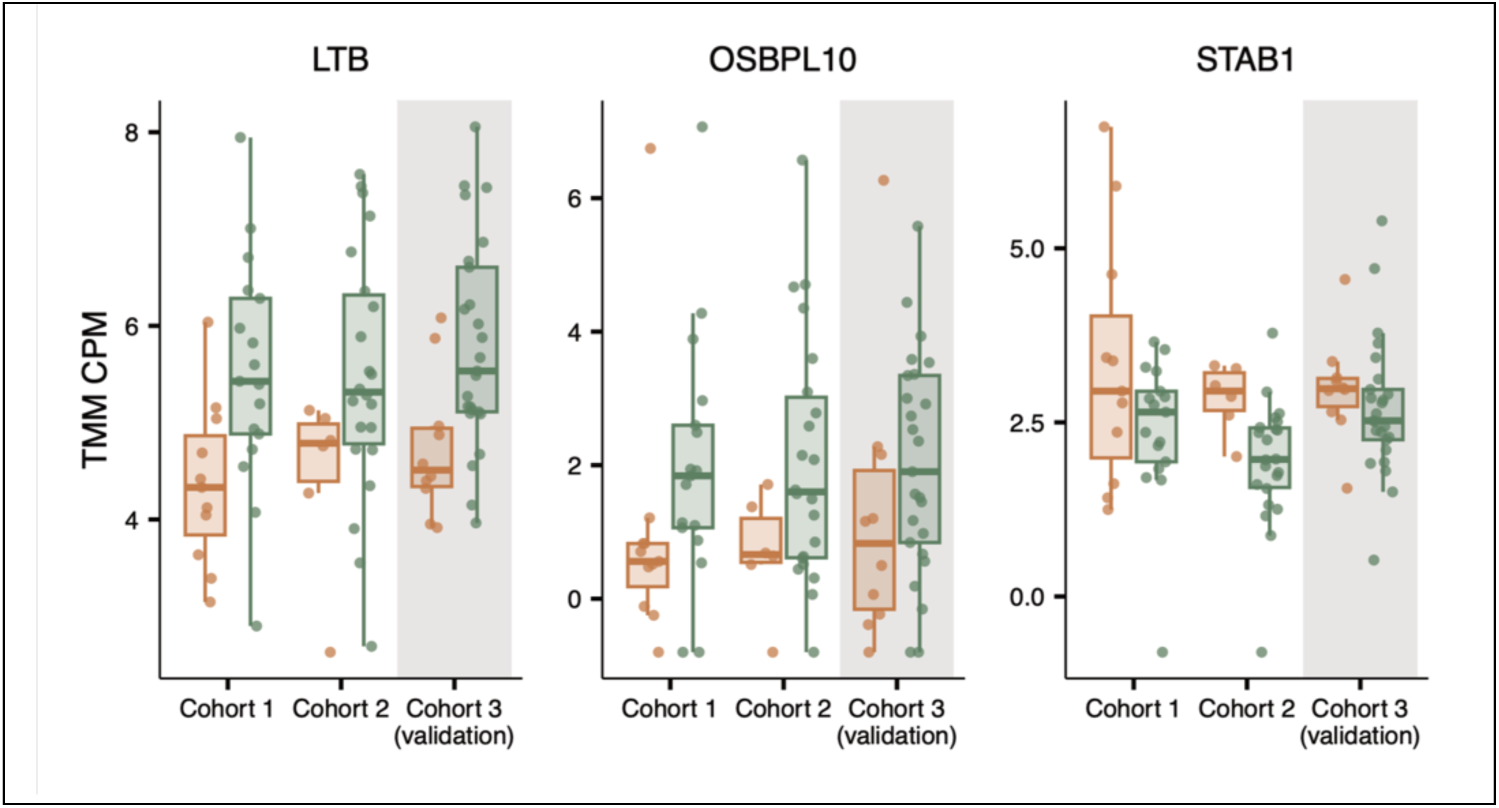
Differentially abundant marker transcripts in cfRNA. Normalized cfRNA abundance (log2 TMM CPM) of *LTB*, *OSBPL10*, and *STAB1* across all three cohorts. Grey shading indicates the validation cohort (Cohort 3).

**Table S1.** Patient demographics.

| Table S1. Patient demographics. |  |  |  |  |  |  |
| --- | --- | --- | --- | --- | --- | --- |
|  |  | Cohort |  |  |  |  |
|  | Total<br>n = 91 (%) | 1<br>n = 28 (31) | 2<br>n = 28 (31) | 3<br>n = 35 (38) |  | p-value |
| **Age at infusion** |  |  |  |  |  |  |
| Median (range) | 65 (24 - 83) | 66 (33 - 83) | 64 (39 - 76) | 65 (24 - 77) |  | 0.77<sup>^</sup> |
| ≤65 | 49 (54) | 13 (46) | 16 (57) | 20 (57) |  | 0.67<sup>†</sup> |
| >65 | 42 (46) | 15 (54) | 12 (43) | 15 (43) |  |  |
| **Diagnosis** |  |  |  |  |  |  |
| De novo DLBCL | 67 (74) | 11 (39) | 24 (86) | 32 (91) |  | < 0.001<sup>†</sup> |
| FL | 4 (4) | 3 (11) | 1 (4) | - |  |  |
| HGBL | 2 (2) | 2 (7) | - | - |  |  |
| PMBCL | 1 (1) | - | - | 1 (3) |  |  |
| tCLL | 1 (1) | 1 (4) | - | - |  |  |
| tFL | 14 (15) | 9 (32) | 3 (11) | 2 (6) |  |  |
| tMZL | 2 (2) | 2 (7) | - | - |  |  |
| **Subtype** |  |  |  |  |  |  |
| Germinal Center | 42 (46) | 16 (57) | 13 (46) | 13 (37) |  | 0.41<sup>†</sup> |
| Non-GCB | 32 (35) | 8 (29) | 10 (36) | 14 (40) |  |  |
| Missing | 17 (19) | 4 (14) | 5 (18) | 8 (23) |  |  |
| **Stage** |  |  |  |  |  |  |
| I | 5 (5) | - | 1 (4) | 4 (11) |  | 0.0049<sup>†</sup> |
| II | 5 (5) | - | 1 (4) | 4 (11) |  |  |
| III | 16 (18) | 4 (14) | 10 (36) | 2 (6) |  |  |
| IV | 62 (68) | 24 (86) | 16 (57) | 22 (63) |  |  |
| Missing | 3 (3) | - | - | 3 (9) |  |  |
| **CNS lymphoma** |  |  |  |  |  |  |
| No | 47 (52) | 11 (39) | 11 (39) | 25 (71) |  | 0.88<sup>†</sup> |
| Yes | 9 (10) | 1 (4) | 2 (7) | 6 (17) |  |  |
| Missing | 35 (38) | 16 (57) | 15 (54) | 4 (11) |  |  |
| **Double-hit** |  |  |  |  |  |  |
| No | 76 (84) | 20 (71) | 24 (86) | 32 (91) |  | 0.038<sup>†</sup> |
| Yes | 12 (13) | 7 (25) | 4 (14) | 1 (3) |  |  |
| Missing | 3 (3) | 1 (4) | - | 2 (6) |  |  |
| **Triple-hit** |  |  |  |  |  |  |
| No | 85 (93) | 25 (89) | 27 (96) | 33 (94) |  | 0.20<sup>†</sup> |
| Yes | 3 (3) | 2 (7) | 1 (4) | - |  |  |
| Missing | 3 (3) | 1 (4) | - | 2 (6) |  |  |
| **ECOG at diagnosis** |  |  |  |  |  |  |
| 0 | 24 (26) | 6 (21) | 7 (25) | 11 (31) |  | 0.52<sup>†</sup> |
| 1 | 41 (45) | 15 (54) | 14 (50) | 12 (34) |  |  |
| 2 | 8 (9) | 1 (4) | 3 (11) | 4 (11) |  |  |
| Missing | 18 (20) | 6 (21) | 4 (14) | 8 (23) |  |  |
| **IPI** |  |  |  |  |  |  |
| 0 | 1 (1) | - | 1 (4) | - |  | 0.034<sup>†</sup> |

|  |  | Cohort |  |  |  |
| --- | --- | --- | --- | --- | --- |
|  | Total<br>n = 91 (%) | 1<br>n = 28 (31) | 2<br>n = 28 (31) | 3<br>n = 35 (38) | p-value |
| 1 | 10 (11) | 1 (4) | 3 (11) | 6 (17) |  |
| 2 | 15 (16) | 2 (7) | 6 (21) | 7 (20) |  |
| 3 | 26 (29) | 15 (54) | 7 (25) | 4 (11) |  |
| 4 | 23 (25) | 5 (18) | 10 (36) | 8 (23) |  |
| 5 | 4 (4) | 1 (4) | 1 (4) | 2 (6) |  |
| Missing | 12 (13) | 4 (14) | - | 8 (23) |  |
| **Prior SCT** |  |  |  |  |  |
| Allo | 1 (1) | 1 (4) | - | - | 0.17* |
| Auto | 22 (24) | 7 (25) | 10 (36) | 5 (14) |  |
| Neither | 66 (73) | 20 (71) | 18 (64) | 28 (80) |  |
| Missing | 2 (2) | - | - | 2 (6) |  |
| **Prior chemotherapy** |  |  |  |  |  |
| No | 9 (10) | 4 (14) | - | 5 (14) | 0.11* |
| Yes | 82 (90) | 24 (86) | 28 (100) | 30 (86) |  |
| **No. of pre-infusion therapies** |  |  |  |  |  |
| Median (range) | 3 (1 - 10) | 3 (1 - 10) | 3 (1 - 5) | 2 (1 - 5) | 0.026* |
| Missing | 7 (8) | 2 (7) | - | 5 (14) |  |
| **Best Response Rate** |  |  |  |  |  |
| CR | 75 (82) | 21 (75) | 24 (86) | 30 (86) |  |
| PR | 6 (7) | 2 (7) | 1 (4) | 3 (9) |  |
| SD/PD | 10 (10) | 5 (18) | 3 (11) | 2 (6) |  |
| **CRS** |  |  |  |  |  |
| Any grade | 81 (89) | 19 (68) | 28 (100) | 34 (97) |  |
| Grade 1-2 | 79 (87) | 19 (68) | 27 (96) | 33 (94) |  |
| Grade 3+ | 2 (2) | 0 (0) | 1 (4) | 1 (3) |  |
| **ICANS** |  |  |  |  |  |
| Any grade | 54 (59) | 12 (43) | 17 (61) | 25 (71) |  |
| Grade 1-2 | 26 (29) | 6 (21) | 10 (36) | 10 (29) |  |
| Grade 3+ | 28 (31) | 6 (21) | 7 (25) | 15 (43) |  |
| *Kruskal-Wallis rank-sum test, *Fisher's exact test |  |  |  |  |  |

